# The association between COVID-19-imposed lockdowns and online searches for toothache using Google Trends

**DOI:** 10.1101/2020.08.01.20157065

**Authors:** Ahmad Sofi-Mahmudi, Erfan Shamsoddin, Peyman Ghasemi, Mona Nasser, Bita Mesgarpour

## Abstract

**Objective:** To assess the association between the lockdowns due to COVID-19 and global online searches for toothache using Google Trends (GT).

**Methods:** We investigated GT online searches for the search terms “toothache” and “tooth pain”, within the past five years. The time frame for data gathering was considered as the initiation and end dates of national/regional lockdowns in each country. Relative search volumes (RSVs) for online Google Search queries in 2019 was considered as the control. We analysed data after normalising based on the Internet penetration rate. We used one-way ANOVA to identify statistical difference for RSVs between 2020 and 2016-2019 for each country. A linear regression model was used to assess whether there is a correlation between RSVs in 2020 and gross domestic production, COVID-19 deaths, dentists’ density, YLDs of oral conditions, Internet access, lockdown duration, Education Index, and dental expenditure per capita.

**Results:** The results of worldwide RSVs for toothache and tooth pain also showed significantly higher values in 2020 compared to the previous four years. Of 23 included countries in our study, 16 showed significantly increased RSVs during the lockdown period compared to the same periods in the past four years. There was a statistically significant relationship between difference of RSVs means in 2020 and in 2016-2019 combined with percent of urban residency (B=-1.82; 95% CI: (-3.38, −0.26); *p*=0.026) and dental expenditure per capita (B=-0.42; 95% CI: (-0.80, −0.05); *p*=0.031) (R^2^=0.66).

**Conclusion:** Generally, the interest in toothache and tooth pain has significantly increased in 2020 compared to the last four years. This could implicitly reinforce the importance of dental care, as urgent medical care worldwide. Governments’ expenditure on oral healthcare and the rate of urban residency, could be mentioned as important factors to direct general populations’ online care-seeking behaviour with regard to dental pain.

## Introduction

Since December 2019, a soaring pandemic has paved its way through the world, the coronavirus disease 2019 (COVID-19). Nearly one month later, the disease was announced by the World Health Organization (WHO) as a public health emergency of international concern (Guo et al. 2020; Wu et al. 2020). Devastating consequences of CVODI-19 go beyond clinical symptoms, and other deteriorations have been consequently addressed in the literature. These could be of many different types, namely financial crisis (Nicola et al. 2020), difficulties in conducting research (Sharpless 2020; Staniscuaski et al. 2020), behavioural alterations (Bavel et al. 2020), management policies (Weible et al. 2020), and healthcare provision problems (Tanne et al. 2020). Accordingly, an effective response to the outbreak and resolving every probable uncertainty related to COVID-19 could only be achieved by scrutinising as comprehensive as possible.

Oral health has shown to be correlated with different aspects of health. Dental problems can not only affect the quality of life, but they can also disrupt people’s individual and/or social life (Baiju et al. 2017; Hescot 2017; Spanemberg et al. 2019; Zucoloto et al. 2016). Accordingly, immediate access to a reliable pain relief source seems to be necessary. Toothache was presented as the most common chief complaint by a study in India in 2015, which was reported by 33% of their participants (1014 patients) (Maheswaran et al. 2015).

Data from studies through the COVID-19 pandemic are also in line with the previous literature before the COVID-19 outbreak with regard to the importance of tooth pain and toothache symptoms. The most common diagnoses in pediatric patients in an Urgent Dental Care Centre in the North East of England and North Cumbria has been reported as irreversible pulpitis and dental trauma (Simpsons et al. 2020). Another study by Carter et al. stated acute pulpal and periapical complaints as the most attendances (for adults and pediatric patients) in urgent dental care centres of Newcastle Dental Hospital (Carter et al. 2020). In another service provision evaluative study, the most frequent diagnosis in a tertiary hospital-based in the UK’s epicentre of the COVID-19 pandemic was reported as irreversible pulpal/apical pathology (63.4% of all diagnoses) (Grossman et al. 2020b). This can be of even more concern when people could not easily access their dental practitioners due to COVID-19 related lockdowns. In several countries, dental offices have been closed or limited due to pandemic and in some countries triage system has been put in place to reduce aerosol generating procedures that resulted that some patients were not able to see dentists when they needed to or were offered less favourable procedures for the problems. Proposing several clinical guidelines and restrictive protocols by major regulatory organizations of the dental medicine might be another harbinger of limited access to dentists during this outbreak ((CDC) 2020; Association 2020). Even in communities that these restriction were not implemented, restriction in public transports limited the possibility for individuals to access dental care. Our hypothesis was that the occurrence of COVID-19 pandemic has substantially afflicted accessibility to the dental discipline.

Real-time data sources are the only fitting mean to address the high-speed alterations in COVID-19-imposed issues in the public health context. Google Trends (GT) has already been proven as a powerful pubic surveillance tool for epidemiological studies (Nuti et al. 2014b; Yeh and Yeh 2019). It is also stated that the more broad target audience of a common disease gets, the more reliable results of GT data records get (Cervellin et al. 2017). Obtaining the most comprehensive results is an essential step while searching through GT service. Accordingly, the terms “toothache” and “tooth pain” were considered as acceptable keywords to assess the online GT search queries related to oral healthcare.

This study sought to assess the association between the lockdowns due to COVID-19 and global online searches for toothache using GT.

## Materials and methods

### Google Trends (GT)

We used GT to investigate internet search activity during the lockdowns due to COVID-19 pandemic. This web service determines the proportion of searches for a user-specified term among all searches performed on Google Search. It then provides a relative search volume (RSV), which is the query share of a particular term for a given location and time period, normalised by the highest query share of that term over the time-series and presented on a scale from 0 to 100 (Nuti et al. 2014a). RSV is presented as “Interest” value in GT website.

### Countries

Inclusion criteria for countries were as follows:

- more than 100 deaths per million due to COVID-19 (till July 14^th^ 2020, data retrieved from: worldometers.info/coronavirus);
- a population more than one million;
- and higher than 50% access to the Internet.

The rationale for our inclusion criteria was that GT does not always have enough total Google Search queries, which depend on population and Internet access. Furthermore, countries with more deaths due to COVID-19 had more strictly measured. We found 24 countries with these characteristics. We used “toothache” and “tooth pain” as the primary search terms for all 24 countries as well as their translation into widely spoken languages for each country using Google Translate, other online dictionaries, and Wikipedia page for “toothache”. To define a cutoff for choosing widely spoken languages in each country, we considered the extent of RSVs in each language which was deemed to reach a minimum level to produce meaningful results in GT. Since the maximum search topics in the GT are predetermined to five slots, the least percentage to cover all the official languages in each country was set as 20%. So, a widely spoken language was considered as a language spoken by more than 20% of the population of that country.

Any query can be explored in GT either as a “search term” or a “topic”. Topics are considered to be more general in terms of searching where they will include all the relevant searched-for keywords. Search terms, on the other hand, can direct the GT results in a more controlled pathway. We gave all search terms as “search term” in Google Trend’s search box except for the worldwide search that we used “toothache” as the disease with “tooth pain” as the search term. Searching for “toothache” as the disease ensured that our online search involved all variations and translations for the query. Full details of search terms we used for each country are illustrated in Appendix 1.

As we gave GT more than one search term for each country, it passed a table with the results of RSV for two or more search terms. One of these search terms in a certain week was considered as the highest RSV (100) and this search term and others for every requested week were considered as a proportion of the highest-RSV-week, from 0 to 100. All RSVs were summed for each week and then normalised by the highest summed RSV.

We gathered RSV data for each country for the past five years and then segregated it from the day of national lockdown due to COVID-19 into three consequent months (12 weeks), using Wikipedia (Wikipedia 2020). For the “Worldwide” searches and countries in which there were no national lockdown dates announced, we used 3/15/2020 and 5/17/2020 as the staring and ending weeks, respectively. We did the same for a similar period of time of 2016-2019 and chose the past year (2019) as the control event.

### Statistical analysis

As Internet access have been probably changing through the past five years, we normalised the data based on the reported Internet penetration rate of each country from 2016-2020. We plotted the trend of “toothache” for each country from five years ago to date. Then a one-way ANOVA statistical test was performed to identify whether there was a statistical difference for RSV scores between the year 2020 and 2016-2019 for each country. Then we used linear regression model to estimate whether there is a correlation between RSV in 2020 and gross domestic production (GDP) per capita (data retrieved from: data.worldbank.org), deaths/million (till 14^th^ July, 2020, data retrieved from: worldometers.info/coronavirus), dentists’ density per 100,000 (data retrieved from: who.int/gho/health_workforce/dentists_density/en), YLDs of oral conditions in 100,000 (Collaborators et al. 2020), estimated individual Internet access in 2020 (based on World Bank’s data for previous years, available from: https://data.worldbank.org/indicator/IT.NET.USER.ZS), lockdown duration (data retrieved from: en.wikipedia.org/wiki/National_responses_to_the_COVID-19_pandemic), Education Index (data retrieved from: en.wikipedia.org/wiki/Education_Index), and dental expenditure per capita (Righolt et al. 2018). Statistical analyses were done using Python v3.6.7 (2018-10-20) (Python Software Foundation, Delaware, United States. http://www.python.org) on Google Colab. A p-value<0.05 was considered as statistically significant.

## Results

The search for our desirable terms returned results for all countries except for Moldova so the analysis included worldwide and 23 countries. Belgium and Romania had the highest and the lowest deaths/million, respectively. More than half of these nations were categorised as high sustainable development index (SDI) countries. Only Sweden had not introduced any lockdowns to date, and other countries had lockdowns with different extents (city (1), state (3), national (19)) and strategies (complete lockdown (19), restriction (2), intelligent lockdown(2)). The median of the start and end date of lockdown were 2020-03-19 and 2020-05-10, respectively. Full details of the characteristics of the included countries are shown in Table 1.

**Table 1.** Characteristics of the included countries

| Country | SDI | Deaths/1M | LD start date | LD end date | LD duration | LD scope | YLDs for oral conditions |
| --- | --- | --- | --- | --- | --- | --- | --- |
| **Belgium** | High | 844 | 2020-03-18 | 2020-05-04 | 48 | National | 288.8 |
| **UK** | High | 660 | 2020-03-23 | 2020-07-04 | 104 | National | 234.3 |
| **Spain** | High-middle | 608 | 2020-03-14 | 2020-05-09 | 57 | National | 201.3 |
| **Italy** | High | 578 | 2020-03-09 | 2020-05-18 | 71 | National | 182.2 |
| **Sweden** | High | 548 | - | - | 0 | - | 246.1 |
| **France** | High | 460 | 2020-03-17 | 2020-05-11 | 56 | National | 210.8 |
| **USA** | High | 418 | 2020-03-19 |  | 118 | State | 213.3 |
| **Chile** | High-middle | 367 | 2020-03-19 |  | 118 | City | 278.1 |
| **Netherlands** | High | 358 | 2020-03-13 |  | 124 | National | 271.1 |
| **Ireland** | High | 353 | 2020-03-12 | 2020-05-18 | 68 | National | 270.4 |
| **Brazil** | Middle | 343 | 2020-03-17 | 2020-05-10 | 55 | State | 345.5 |
| **Ecuador** | High-middle | 287 | 2020-03-16 | 2020-03-31 | 16 | National | 328.2 |
| **Mexico** | High-middle | 275 | 2020-03-23 | 2020-06-01 | 71 | National | 339.5 |
| **Canada** | High | 233 | 2020-03-17 | 2020-05-04 | 49 | State | 217.8 |
| **Switzerland** | High | 227 | 2020-03-20 | 2020-04-26 | 37 | National | 218.0 |
| **Armenia** | High-middle | 196 | 2020-03-24 | 2020-05-04 | 41 | National | 344.5 |
| **North Macedonia** | High-middle | 185 | 2020-03-21 | 2020-05-26 | 66 | National | 247.6 |
| **Portugal** | High-middle | 163 | 2020-03-19 | 2020-04-02 | 14 | National | 234.0 |
| **Iran** | Middle | 157 | 2020-03-14 | 2020-04-20 | 37 | National | 228.9 |
| **Germany** | High | 109 | 2020-03-23 | 2020-05-10 | 48 | National | 248.1 |
| **Colombia** | High-middle | 107 | 2020-03-25 | 2020-06-30 | 97 | National | 337.5 |
| **Denmark** | High | 105 | 2020-03-13 | 2020-04-13 | 31 | National | 293.3 |
| **Romania** | High-middle | 100 | 2020-03-25 | 2020-05-12 | 48 | National | 261.4 |

There were no significant differences between RSVs for countries with and without national lockdown (P=0.694). When comparing 2020 with other previous four years combined, 16 countries and the world showed statistically significant higher RSV values. However, Colombia, Chile, Ireland, and Portugal had not significantly higher RSVs in 2020 compared to every past four years (Table 2). Sweden, Netherlands, Ecuador, and North Macedonia had lower 2020 RSVs comparing to previous years (Table 3). The worldwide RSV trend in three months post lockdown for 2020-2016 is illustrated in Figure 1 (Appendix 2 for countries trends) and RSVs trend in the past five years is shown in Figure 2.

**Table 2.** ANOVA results

| Country | 2020 with 2019-6 |  | 2020 with 2019 |  | 2020 with 2018 |  | 2020 with 2017 |  | 2020 with 2016 |  |
| --- | --- | --- | --- | --- | --- | --- | --- | --- | --- | --- |
|  | F-value | P-value | F-value | P-value | F-value | P-value | F-value | P-value | F-value | P-value |
| **World** | 3243.9 | **<0.001** | 2380.5 | **<0.001** | 2565.8 | **<0.001** | 2948.2 | **<0.001** | 3328.2 | **<0.001** |
| **Belgium** | 3.9 | 0.053 | 1.8 | 0.194 | 1.2 | 0.289 | 2.5 | 0.129 | 5.2 | **0.033** |
| **UK** | 130.7 | **<0.001** | 55.5 | **<0.001** | 63.2 | **<0.001** | 105.8 | **<0.001** | 131.9 | **<0.001** |
| **Spain** | 54.0 | **<0.001** | 31.3 | **<0.001** | 27.5 | **<0.001** | 31.0 | **<0.001** | 12.2 | **0.002** |
| **Italy** | 204.0 | **<0.001** | 76.1 | **<0.001** | 70.2 | **<0.001** | 74.9 | **<0.001** | 85.4 | **<0.001** |
| **Sweden** | 0.4 | 0.519 | 0.1 | 0.704 | 3.9 | 0.059 | 0.2 | 0.640 | <0.1 | 0.878 |
| **France** | 112.4 | **<0.001** | 55.5 | **<0.001** | 56.0 | **<0.001** | 84.3 | **<0.001** | 41.5 | **<0.001** |
| **USA** | 120.2 | **<0.001** | 62.8 | **<0.001** | 72.1 | **<0.001** | 105.3 | **<0.001** | 152.5 | **<0.001** |
| **Chile** | 6.0 | **0.017** | 3.1 | 0.094 | 9.7 | **0.005** | 1.5 | 0.239 | 7.3 | **0.012** |
| **Netherlands** | 0.2 | 0.673 | 0.2 | 0.648 | 1.9 | 0.187 | <0.1 | 0.838 | 0.3 | 0.599 |
| **Ireland** | 7.9 | **0.007** | 2.0 | 0.174 | 3.9 | 0.060 | 4.6 | **0.044** | 6.6 | **0.017** |
| **Brazil** | 429.7 | **<0.001** | 552.2 | **<0.001** | 680.8 | **<0.001** | 739.4 | **<0.001** | 1198.4 | **<0.001** |
| **Ecuador** | 0.6 | 0.431 | <0.1 | 0.915 | 1.2 | 0.280 | <0.1 | 0.812 | 3.6 | **0.007** |
| **Mexico** | 36.9 | **<0.001** | 21.4 | **<0.001** | 22.8 | **<0.001** | 28.2 | **<0.001** | 14.0 | **0.001** |
| **Canada** | 38.0 | **<0.001** | 10.4 | **0.004** | 24.7 | **<0.001** | 23.3 | **<0.001** | 46.4 | **<0.001** |
| **Switzerland** | 2.1 | 0.154 | 1.9 | 0.185 | 3.4 | 0.079 | <0.1 | 0.774 | 2.4 | 0.137 |
| **Armenia** | 26.8 | **<0.001** | 14.7 | **<0.001** | 26.2 | **<0.001** | 13.8 | **<0.001** | 33.3 | **<0.001** |
| **North Macedonia** | 0.2 | 0.688 | <0.1 | 0.918 | 0.7 | 0.413 | 0.3 | 0.602 | 2.4 | 0.134 |
| **Portugal** | 5.4 | **0.023** | 3.9 | 0.062 | 3.4 | 0.078 | 0.4 | 0.534 | 4.9 | **0.037** |
| **Iran** | 99.2 | **<0.001** | 38.7 | **<0.001** | 34.6 | **<0.001** | 50.0 | **<0.001** | 99.0 | **<0.001** |
| **Germany** | 20.0 | **<0.001** | 22.5 | **<0.001** | 14.8 | **<0.001** | 10.9 | **0.003** | 7.8 | **0.010** |
| **Colombia** | 12.0 | **<0.001** | 2.5 | 0.126 | 6.9 | **0.015** | 7.6 | **0.012** | 7.7 | **0.011** |
| **Denmark** | 2.6 | 0.110 | 0.2 | 0.651 | 1.5 | 0.230 | 0.9 | 0.348 | 12.1 | **0.002** |
| **Romania** | 19.8 | **<0.001** | 7.0 | **0.014** | 8.1 | **0.009** | 6.8 | **0.016** | 9.3 | **0.006** |

**Table 3.** Mean RSVs by country for each year

| **Country** | **Mean 2020** | **Mean 2019** | **Mean 2018** | **Mean 2017** | **Mean 2016** |
| --- | --- | --- | --- | --- | --- |
| **World** | 94.4 | 77.8 | 73.6 | 71.9 | 66.3 |
| **Belgium** | 32.4 | 30.2 | 30.5 | 26.8 | 24.4 |
| **UK** | 88.7 | 72 | 68.6 | 63.2 | 59.9 |
| **Spain** | 79.2 | 64.1 | 64.8 | 64.9 | 70.7 |
| **Italy** | 69.8 | 59.1 | 60.8 | 63 | 60.1 |
| **Sweden** | 51.2 | 52.5 | 45.9 | 52.6 | 56.2 |
| **France** | 81.1 | 65.6 | 66.7 | 61.4 | 70.3 |
| **USA** | 87.3 | 83.5 | 82.7 | 78.2 | 71.7 |
| **Chile** | 64.1 | 59.1 | 55.3 | 64.6 | 55.2 |
| **Netherlands** | 55.9 | 54.5 | 62.7 | 55.5 | 58.7 |
| **Ireland** | 69.7 | 67.7 | 63.3 | 61.9 | 61.3 |
| **Brazil** | 92.9 | 74 | 64.7 | 58 | 49.7 |
| **Ecuador** | 21 | 27.1 | 20.2 | 30.1 | 14.8 |
| **Mexico** | 21.2 | 18.3 | 18.5 | 18 | 19.2 |
| **Canada** | 85.5 | 78.5 | 69.8 | 71.6 | 62.8 |
| **Switzerland** | 43.4 | 41.8 | 36.8 | 45.7 | 39.1 |
| **Armenia** | 36.3 | 17 | 12.7 | 18.7 | 8.5 |
| **North Macedonia** | 16.6 | 20.9 | 14 | 25.3 | 9.2 |
| **Portugal** | 43.6 | 39.1 | 39.2 | 48.8 | 37.3 |
| **Iran** | 78.5 | 68.9 | 68.7 | 61.6 | 49.1 |
| **Germany** | 68 | 65.3 | 67 | 68.9 | 69.9 |
| **Colombia** | 23.6 | 23.7 | 16.3 | 16.3 | 16.6 |
| **Denmark** | 34.3 | 32.8 | 29.8 | 30.1 | 23.8 |
| **Romania** | 52.3 | 46.8 | 45.9 | 47.8 | 43.6 |

**Figure 1.**
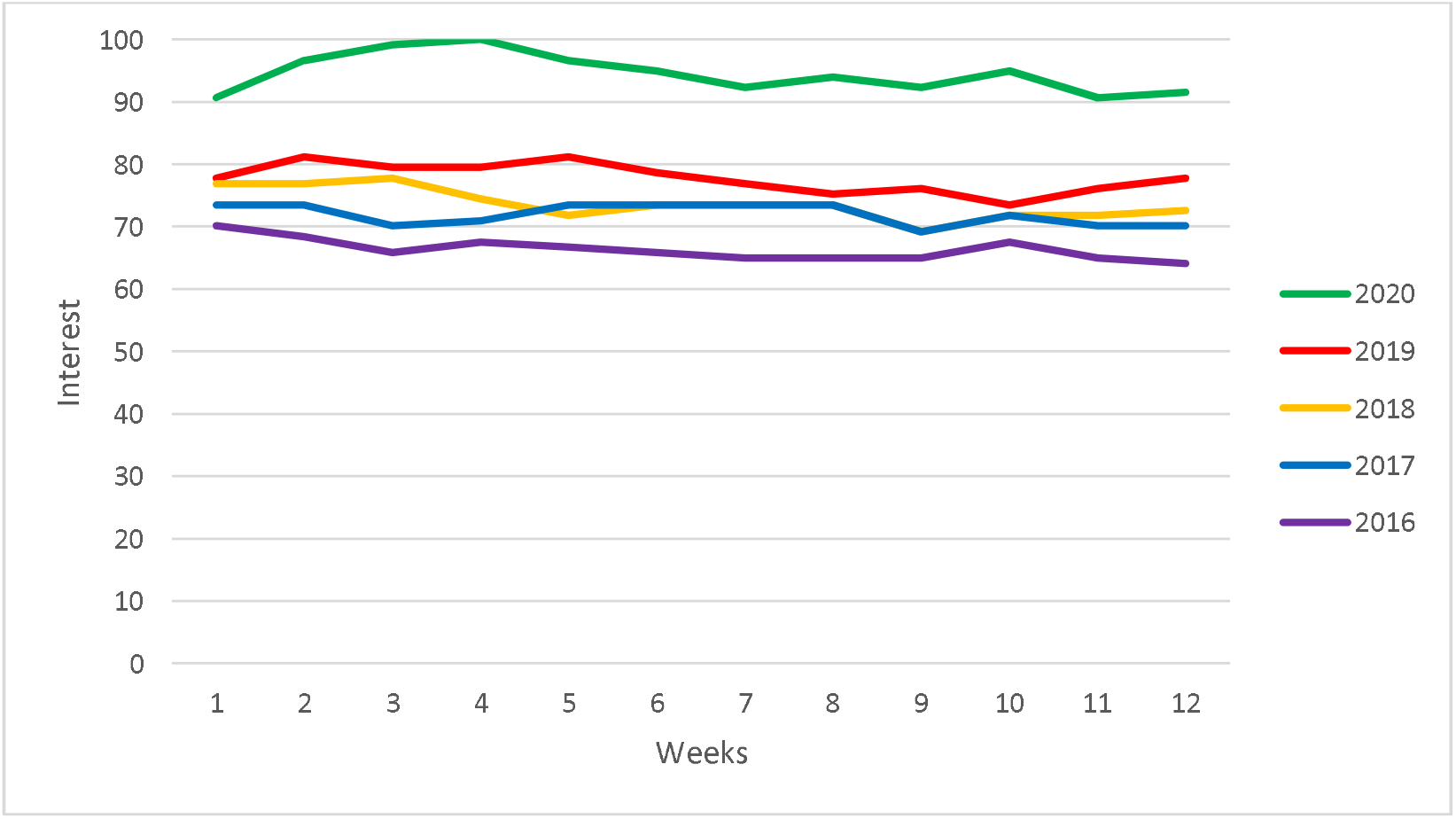
Worldwide RSV trend in three months post lockdown

**Figure 2.**
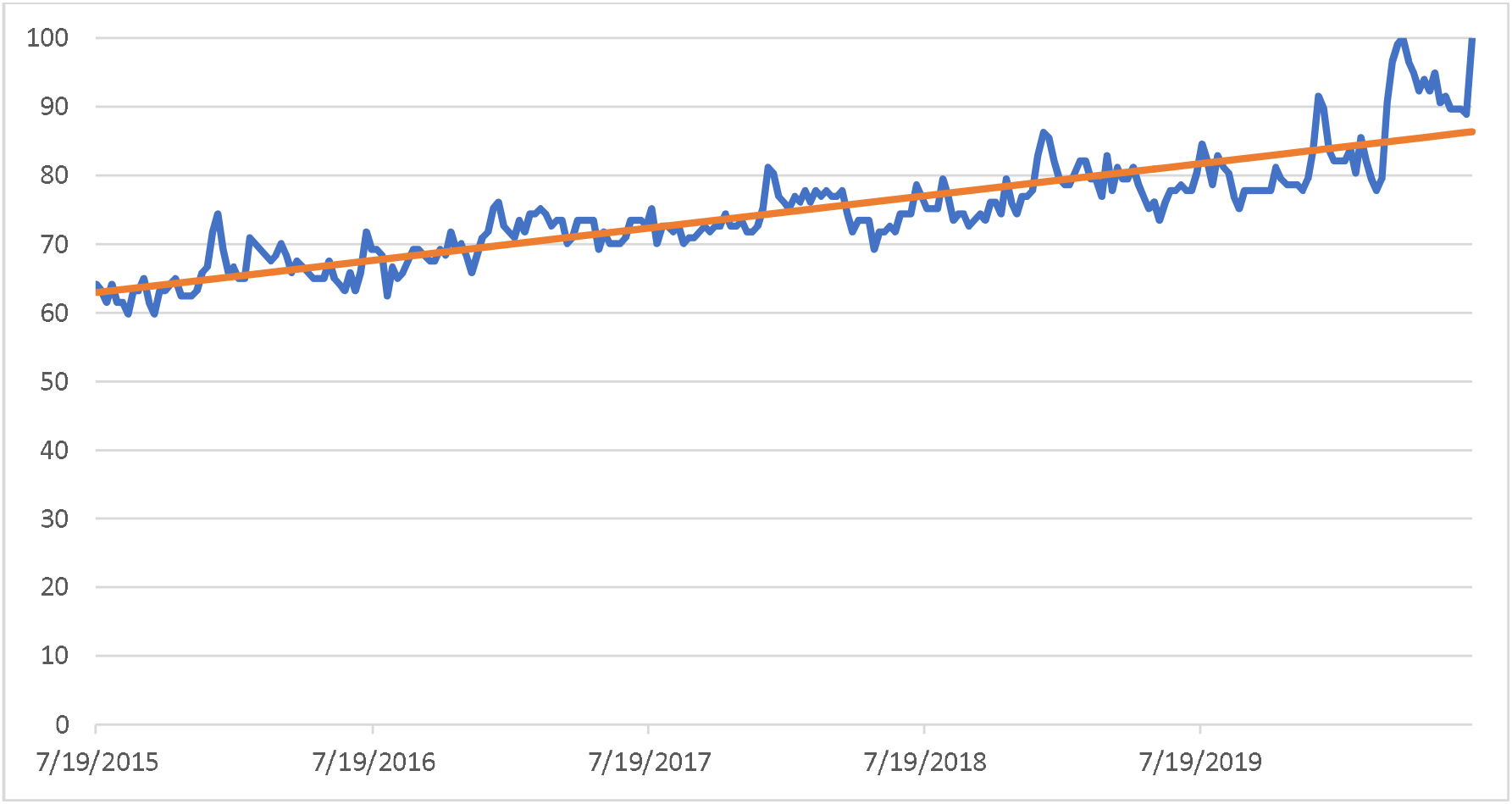
Worldwide RSVs trend for past five years

The linear regression models showed that there is a statistically significant relationship between the difference of RSVs means in 2020 and in 2016-2019 combined with per cent of urban residency (B=-1.82, 95% CI: (-3.38, −0.26), P=0.026) and dental expenditure per capita (B=-0.42, 95% CI: (-0.80, −0.05) P=0.031). The chosen variables explained 66% of the variations between countries. Full details of the linear regression results are shown in Table 4.

**Table 4.** Regression results

|  | **Coefficient (95% CI)** | **P-value** |
| --- | --- | --- |
| **(Constant)** | 0.33 (-0.32, 0.98) | 0.293 |
| **GDP per capita** | -0.27 (-0.73, 0.19) | 0.221 |
| **Deaths per million** | 0.10 (-0.22, 0.43) | 0.495 |
| **YLDs for oral disorders per 100,000** | -0.58 (-1.75, 0.59) | 0.299 |
| **Dentists' density** | 0.38 (-0.01, 0.77) | 0.054 |
| **Estimated individual Internet access at 2020** | 0.71 (-2.99, 4.42) | 0.682 |
| **Urban residency** | **-1.82 (-3.38, -0.26)** | **0.026** |
| **Education Index** | 1.06 (-3.68, 5.81) | 0.634 |
| **Lockdown duration** | 0.14 (-0.21, 0.48) | 0.403 |
| **Dental expenditure per capita** | **-0.42 (-0.80, -0.05)** | **0.031** |
| **R<sup>2</sup>** | **0.66** |  |

## Discussion

Dentistry has been described as being a high-risk medical profession for viral infections, like COVID-19 (Dave et al. 2020; Ge et al. 2020; Shamsoddin 2020). The predicament of following cautious measures and clinical guidelines, behavioural responses to stress, and regulatory restrictions could all be thought of as critical factors decreasing the access to dental services in the time of COVID-19 outbreak. Secondly, dental services (diagnostic or therapeutic) are mainly provided in the private sector in many countries (Brennan et al. 2008; Cascaes et al. 2018; Kiadaliri et al. 2013; Mertz and O’Neil 2002; Widström et al. 2001). This could lead to lower availability of dental services which would get exaggerated during public health emergencies - since many private practitioners may choose not to work during the emergency.

Our study aimed to evaluate the association between COVID-19-imposed lockdowns and global online searches for toothache using GT. Knowing that toothache and tooth pain are the most common symptoms in dental patients and could cause some major complications in habitual and/or working activities, they were considered as the main representatives for dental needs. Among the primary set of 23 countries in our analyses, 16 presented statistically significant higher RSV values (with regard to our keywords) in 2020 compared to the mean RSV in the past 4 years. These results partly reinforce our hypothesis about the major role of COVID-19 pandemic on accessibility to dental service during the government-imposed curfews and lockdowns.

A significant negative correlation was seen between the difference of mean RSVs of 2020 and the last four years, with per cent of urban residency (B=-1.82). Furthermore, a noticeable relationship with the same pattern could be seen between dentists’ density and the RSV difference (B=0.38, P=0.054). These findings suggest that the more accessible dental offices get, the less online google searches about toothache and/or tooth pain will generally get. This implies that care-seeking behaviour (CSB) of the patients could majorly change by the way they deem dental services accessible to them. In Belgium, for instance, with the highest urban residency (98%), people might probably directly contact their dental practitioners upon feeling pain. Another significant correlation was evident between the RSV difference and dental expenditure per capita (DEPC) (B=-0.42). The DEPC here was referring to the financial resources each government has devoted to dental healthcare for each individual. This significant negative relationship implies that the more financial and organizational structures of a country will spend money on the general populations’ dental health, the less public online search will be conducted regarding toothache and/or tooth pain. This could be of even more sophisticated nature given that the previous oral health status of the population and the level of trust that general population have about their healthcare system, could majorly impact their CSB during health emergencies.

Healthier populations who believe more strongly in their healthcare systems’ stewardships, might represent lower RSVs in Google online searches about toothache and/or tooth pain.

Since various restrictive health policies (social distancing, national lockdowns, restrictive professional protocols, etc.) have been applied in response to COVID-19 pandemic at different regions, the results have also been highly heterogeneous. Political position, cultural tendencies, economic resiliency, and healthcare system development have all been mentioned as major role players in determining the reactions to COVID-19 in each country and/or region (Figari and Fiorio 2020; Grasselli et al. 2020; Grossman et al. 2020a; Painter and Qiu 2020).

Lack of significant difference in online search RSVs in 7 countries out of 23 (regarding toothache and tooth pain), could be majorly related to CSB among the population. Whether an individual seeks information from online databases using GT or not could be different among various regions due to several causes. As can be seen, 5 of 7 countries with the non-significant differences were EU members -except for Ecuador and North Macedonia. The mean use of Google Search Engine is reported to be above 95% for all these countries (data from https://gs.statcounter.com/search-engine-market-share). Accordingly, the difference in search engine market share cannot be suggested as a possible cause for this issue.

Media has been shown to affect general online searches for the COVID-19 more than epidemiological data does (Szmuda et al. 2020). Social media and any other popular online platform informing the public about COVID-19 figures could have probably affected the CSB in the general population in each country/region.

Through our literature review, very few similar studies were found which had scrutinized oral health-seeking behaviour of patients. No published study was found determining oral health-seeking behaviour during the COVID-19 pandemic. Nevertheless, one study assessed the Internet search trends of the oral health-seeking behaviour of Filipinos from 2010 to 2020 (Dalanon and Matsuka). According to their results, the most prevalent oral health problems in the Philippines are dental caries and edentulousness. Obtaining these data from a user-friendly data record like GT is utterly emphasized in this study. Another assessment of search Internet search trends for common oral problems was done in 2014 (Harorli and Harorli 2014). The term “toothache” was represented as the most commonly searched query between 2004 to 2014. They also claimed that toothache is the primary source of patient complaints in dentistry. Tao et al. have conducted a study on nature and diffusion of COVID-19 related oral health information on Chinese social media (Tao et al. 2020). They analyzed 15900 tweets regarding oral health and dentistry information from Weibo. Their study showed that the time and geographic distribution of tweets shared similarities with epidemiological data of the COVID-19 outbreak in China.

Tweets containing home oral care and dental services content were the most frequently shared information. Their study emphasized the critical role of governments, regulatory organizations, media, and dental professionals to actively contribute to online social medias, especially throught a public healthcare emergency.

We tried to utilize a comprehensive and pragmatic methodology so that no online search results would be neglected. By considering different spoken languages based on their popularity in each country, we ensured not losing lexical, semantic, and grammatical nuances during translation.

However, GT-derived data should be interpreted cautiously there is no certain way to define the population, which contributed to the data sample. Prompt fluctuations in searching trend can also happen due to media and news agenda (locally or broader aspects). Additionally, it can never be traced if one had searched a term out of curiosity or to address their personal health needs.

Another implicit bias is the Internet accessibility bias which basically refers to disparities of Internet access among different social groups (based on their demographic status, namely literacy level, socio-economic status, age, etc.).

## Conclusion

Significant differences in online search terms relating to dental problems during COVID-19 pandemic compared to the recent years could ultimately express a necessity for a further online contribution of all oral healthcare professionals. Addressing dental healthcare provision as a highly-sought-after medical need would be another critical aspect of this study. Additionally, the government’s expenditures on oral healthcare and the urban residency rate could be considered as essential factors affecting the care-seeking behaviour of the general population regarding toothache and/or tooth pain.

## Data Availability

Data will be available upon request.

## Appendices

Appendix 1. Search terms for each country Reference:

https://www.cia.gov/library/publications/the-world-factbook/fields/402.html

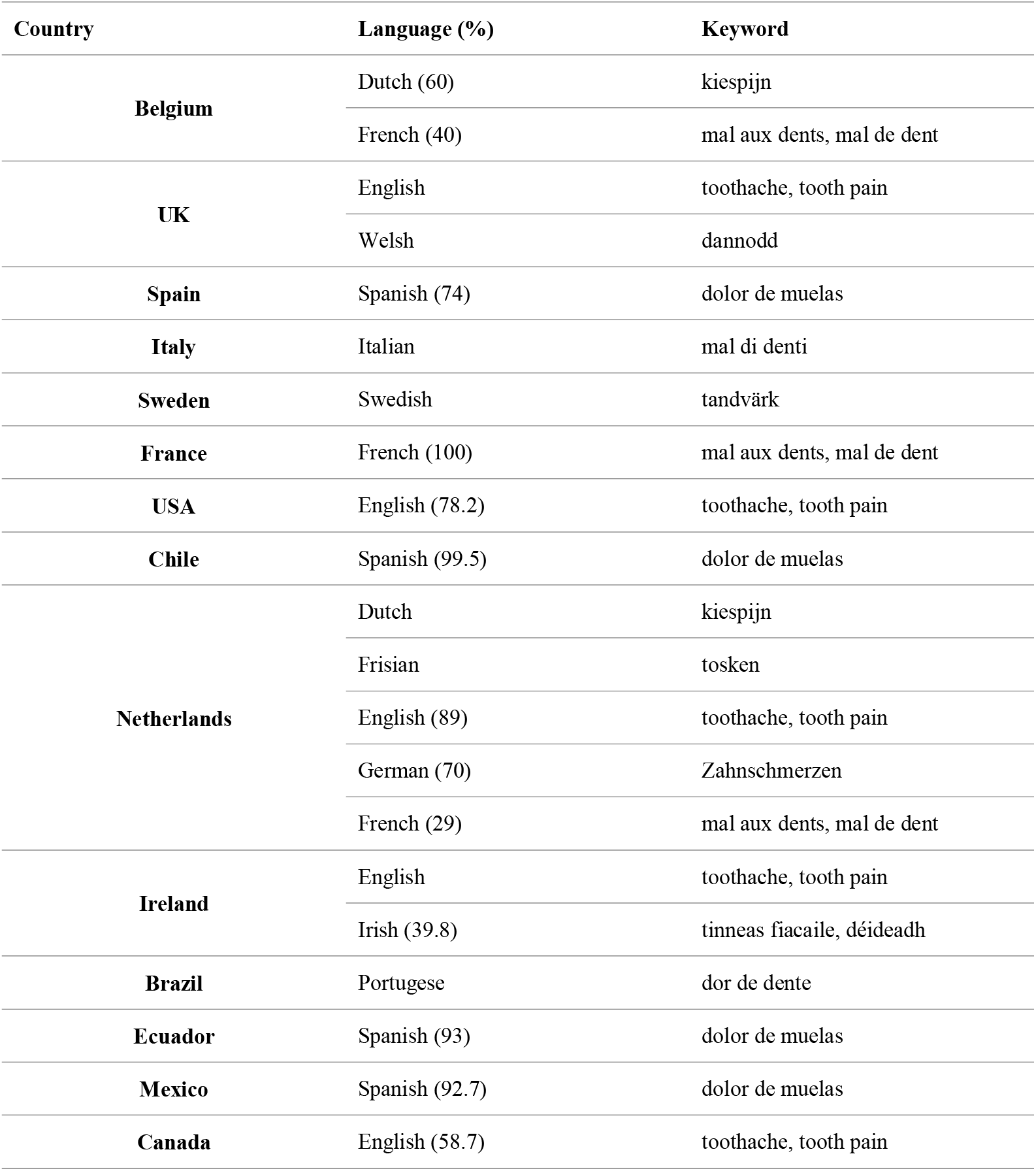
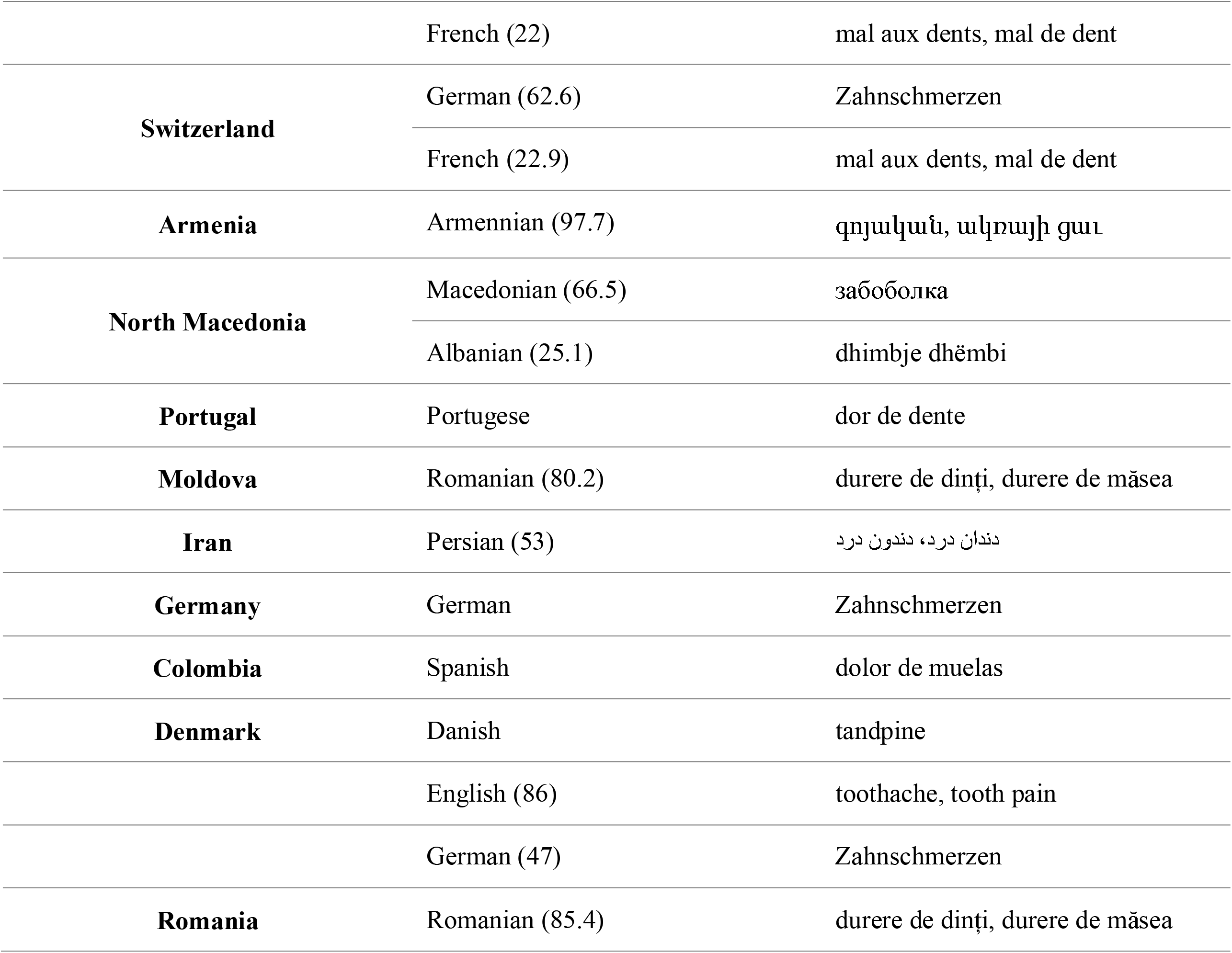

Appendix 2. RSV trends for each country

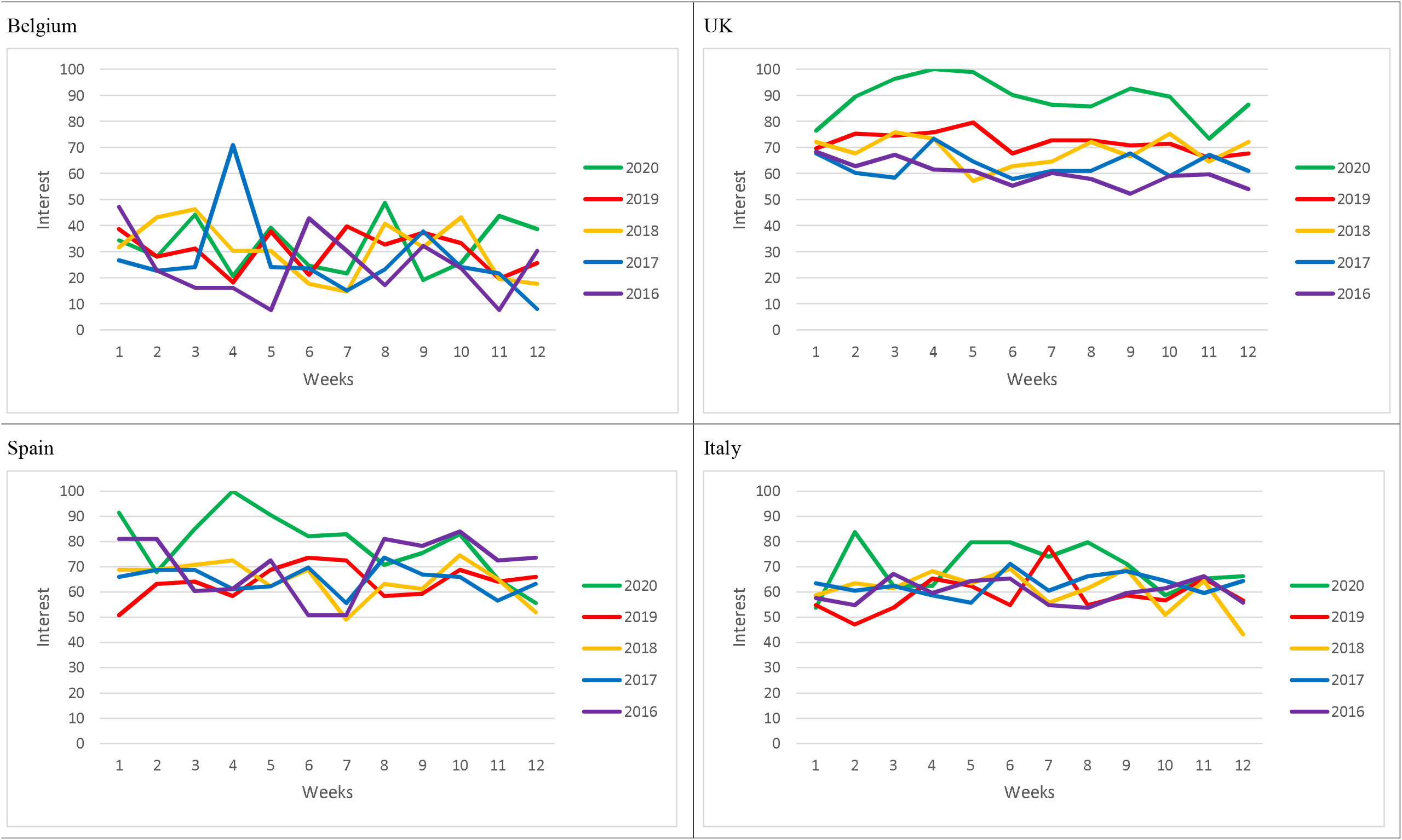
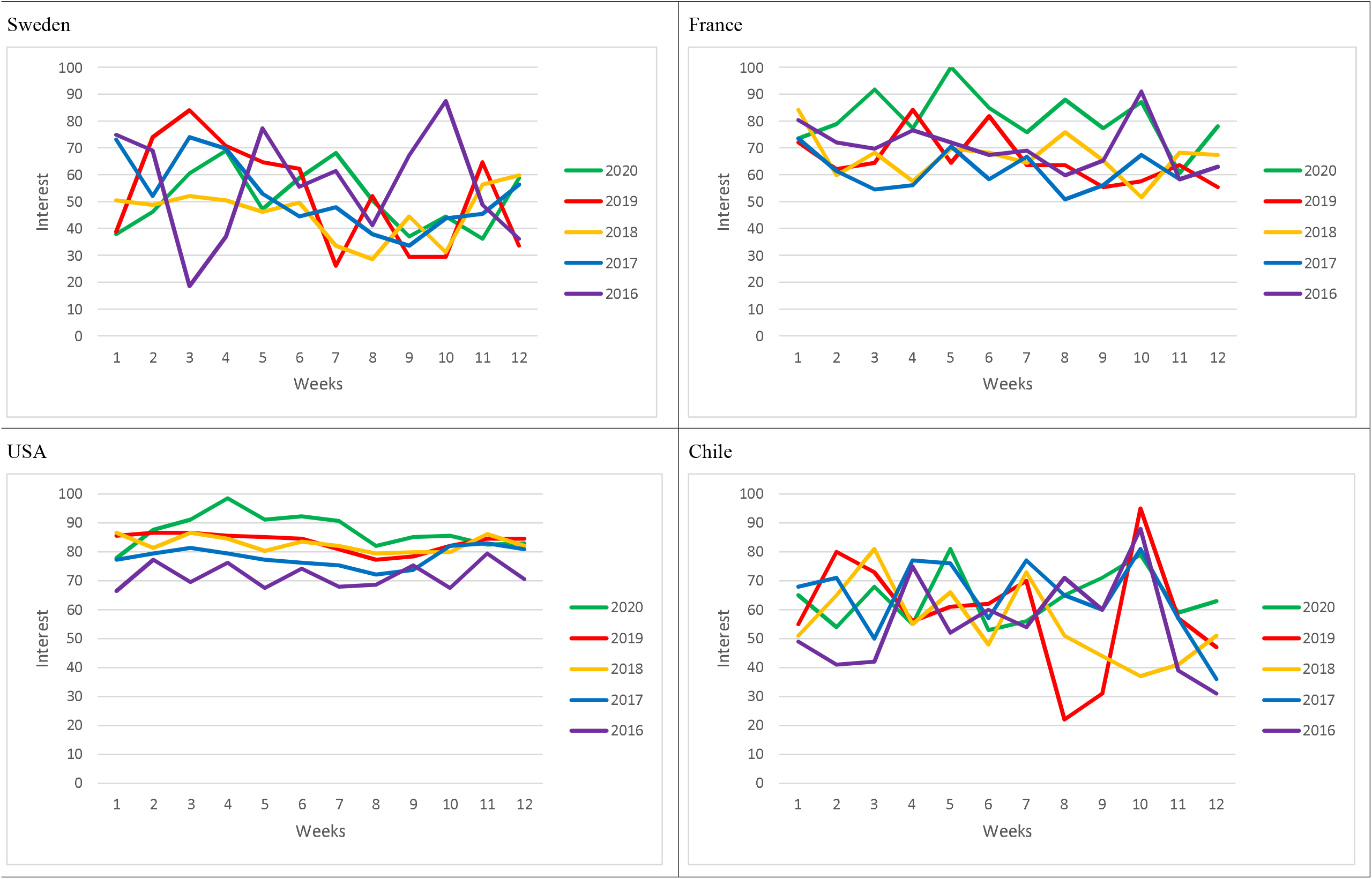
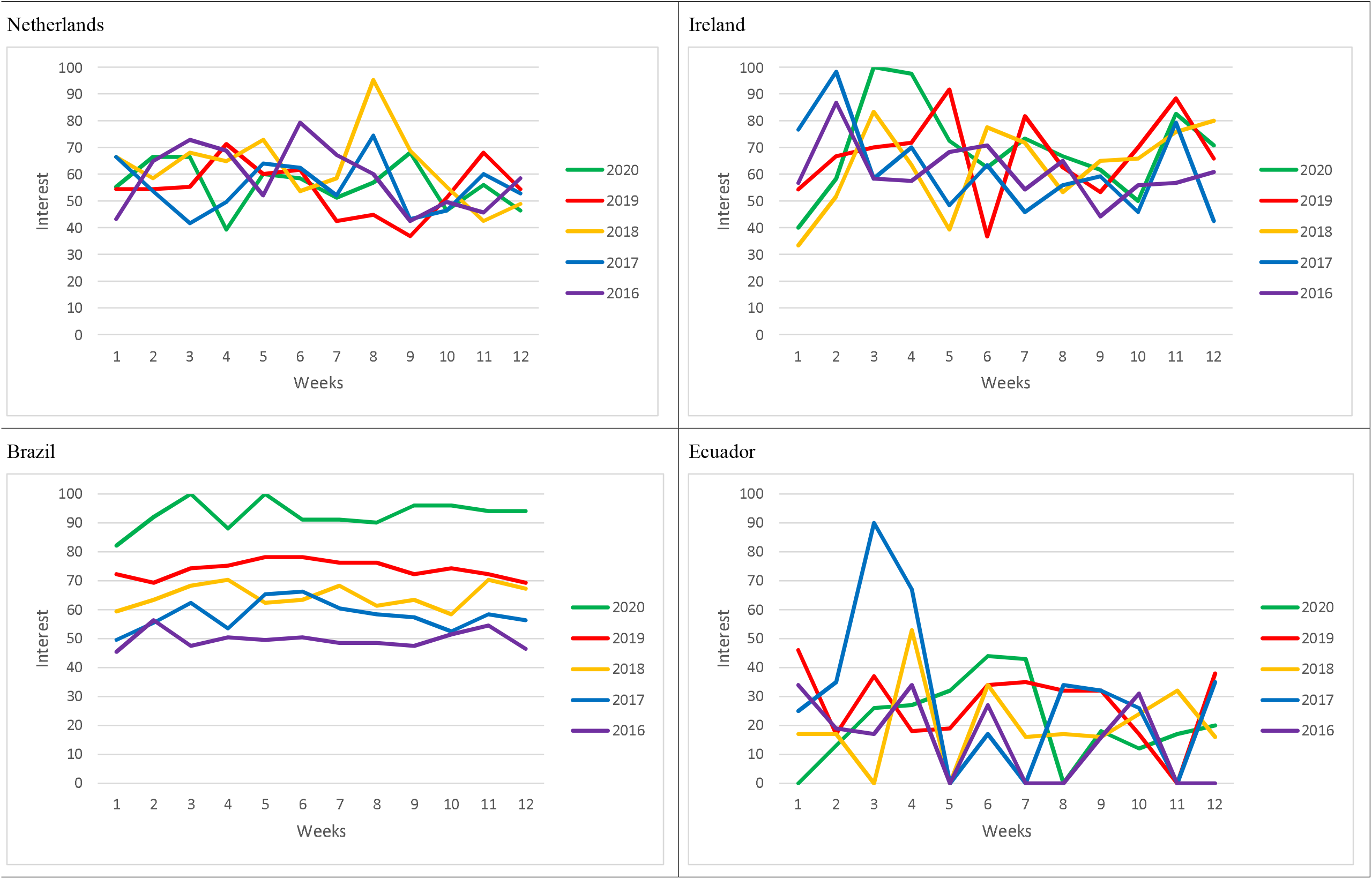
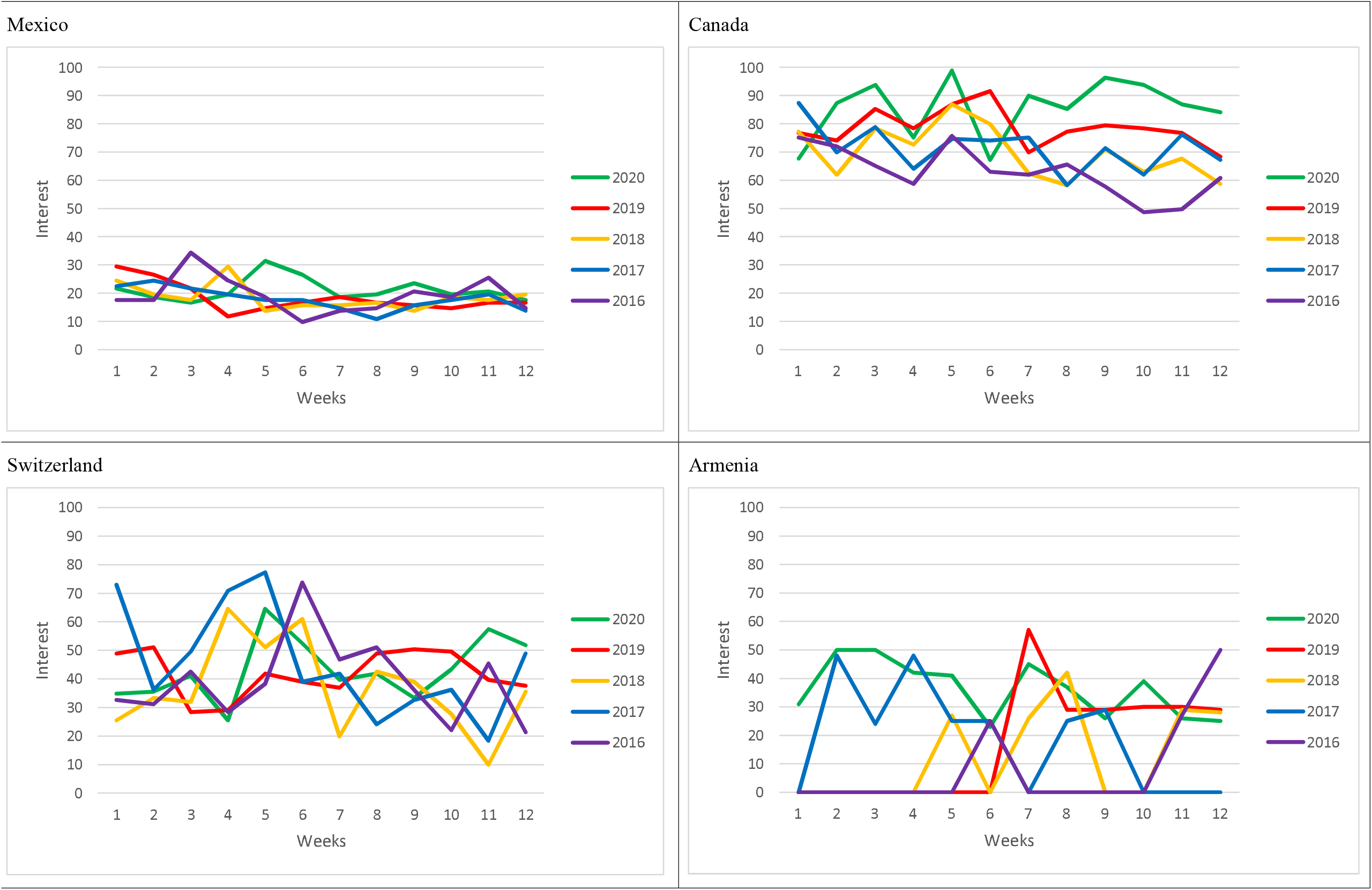
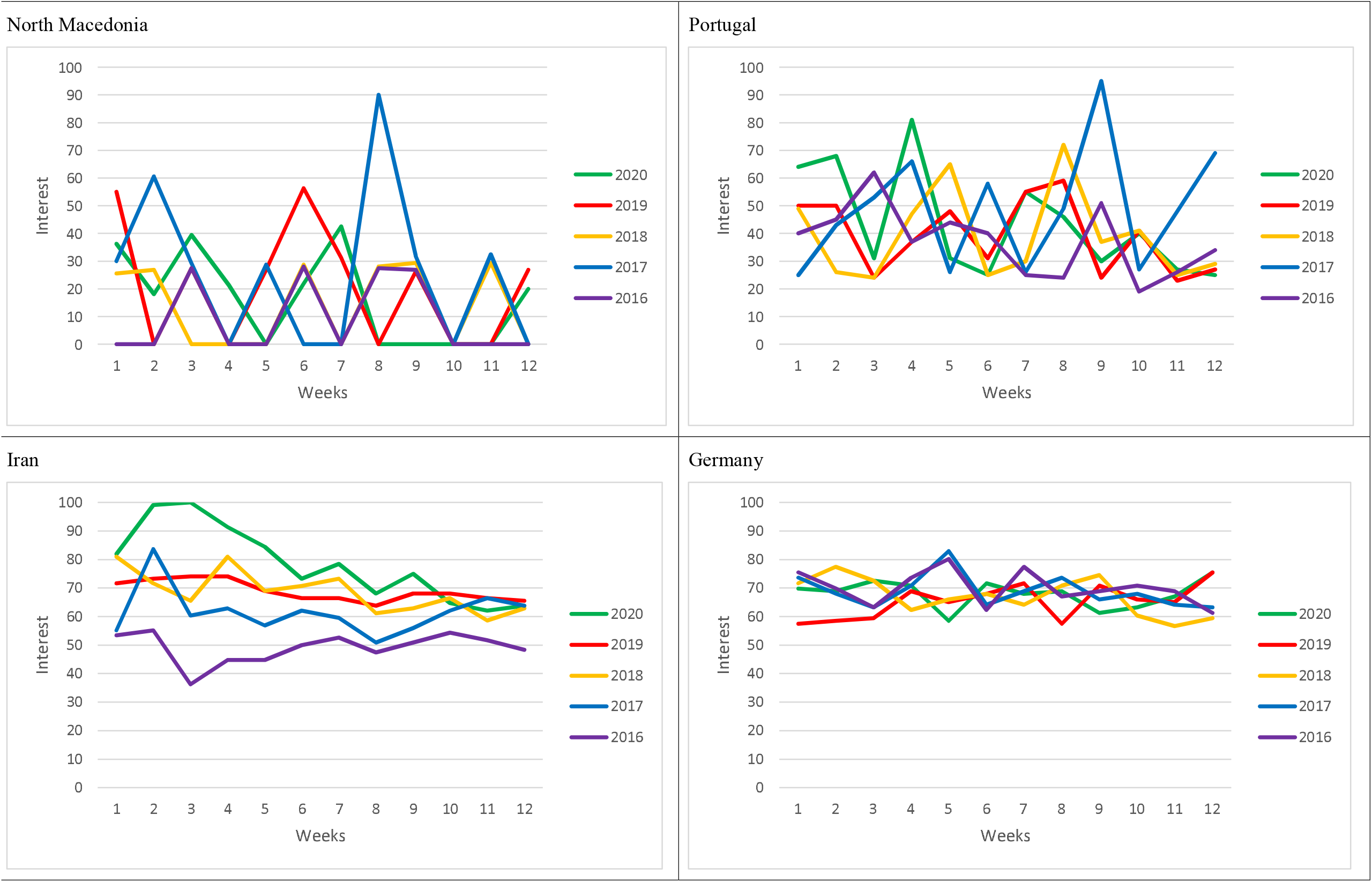
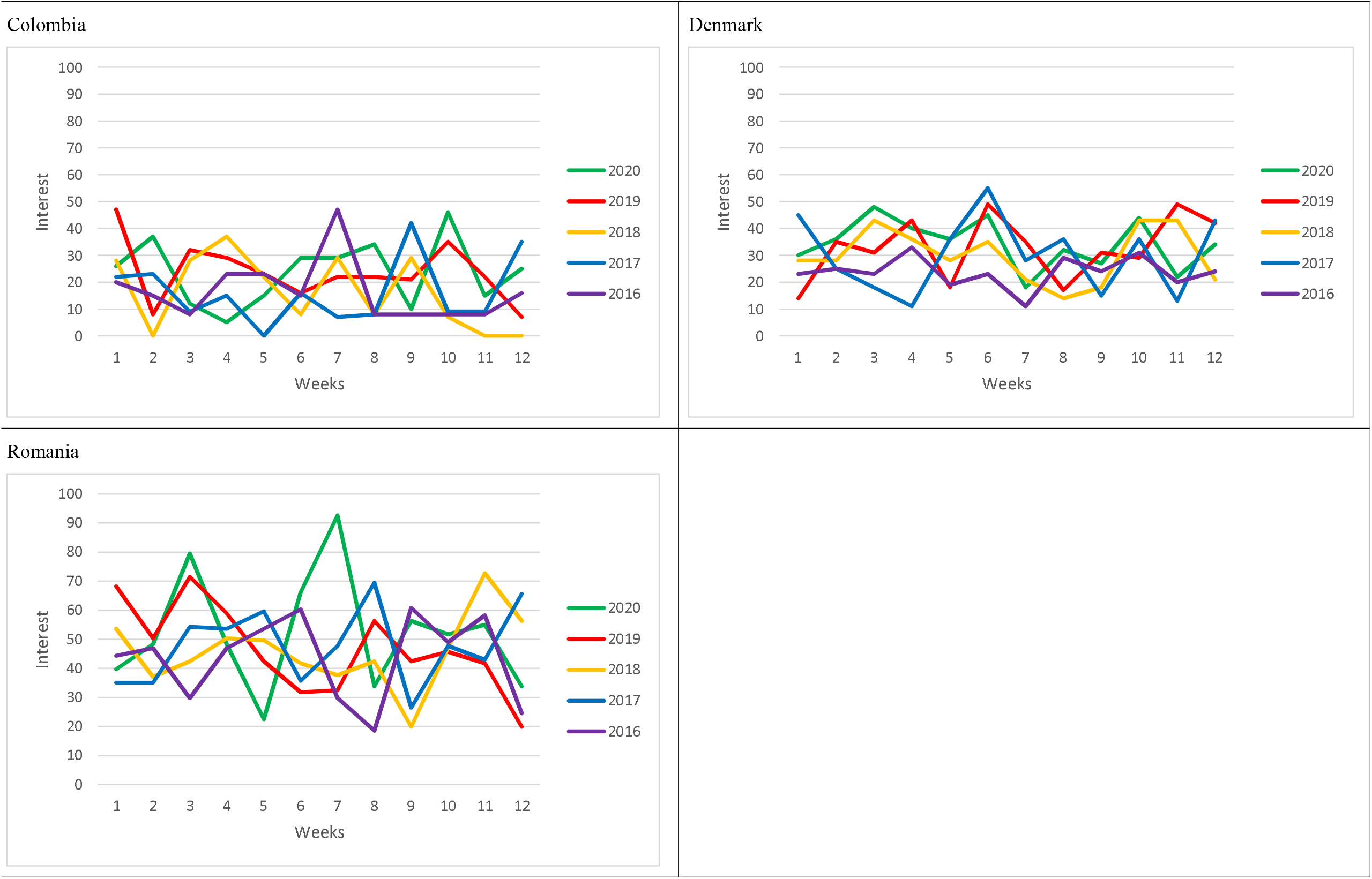

North Macedonia Portugal

Appendix 3. RSVs trends for past five years

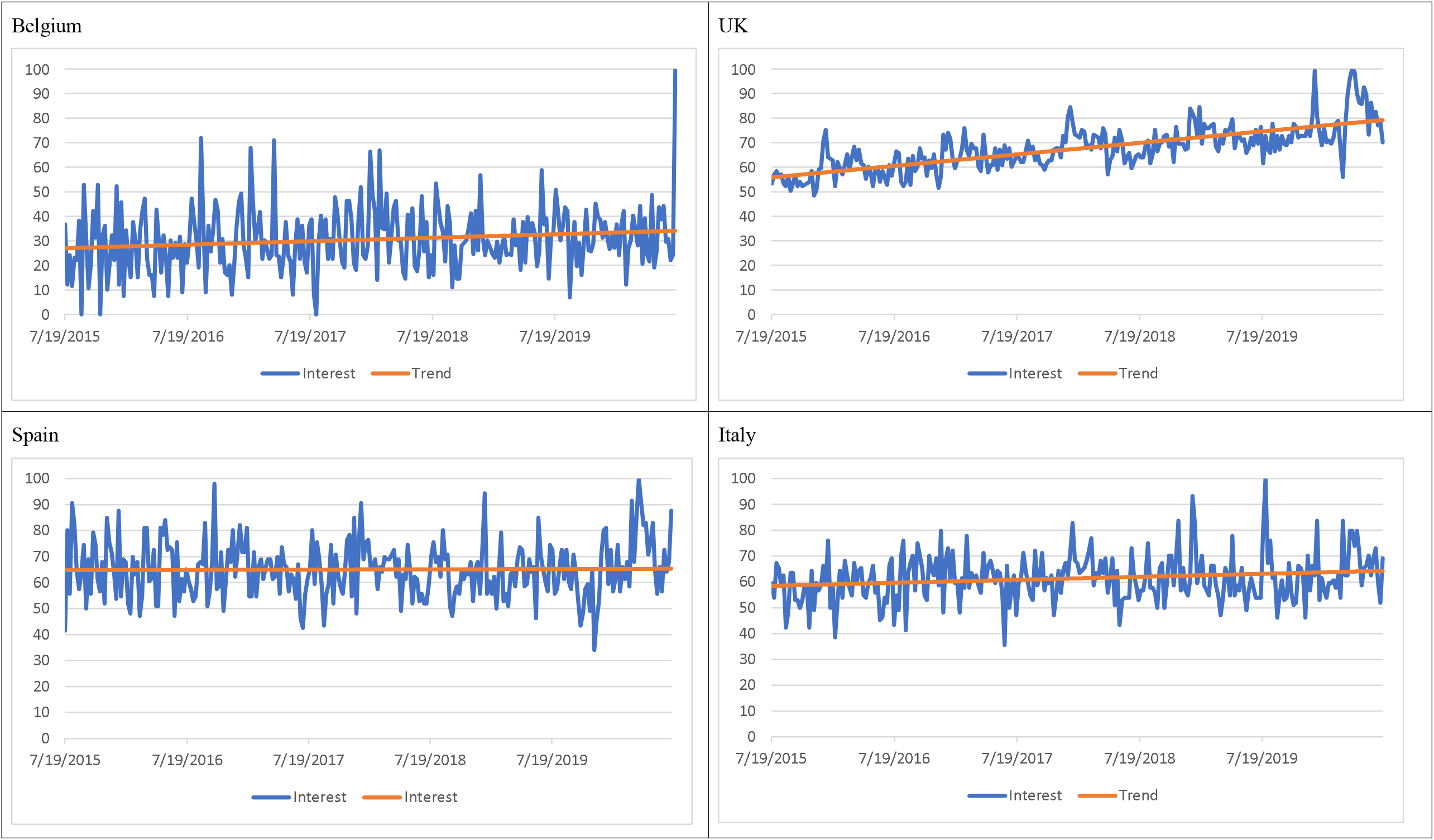
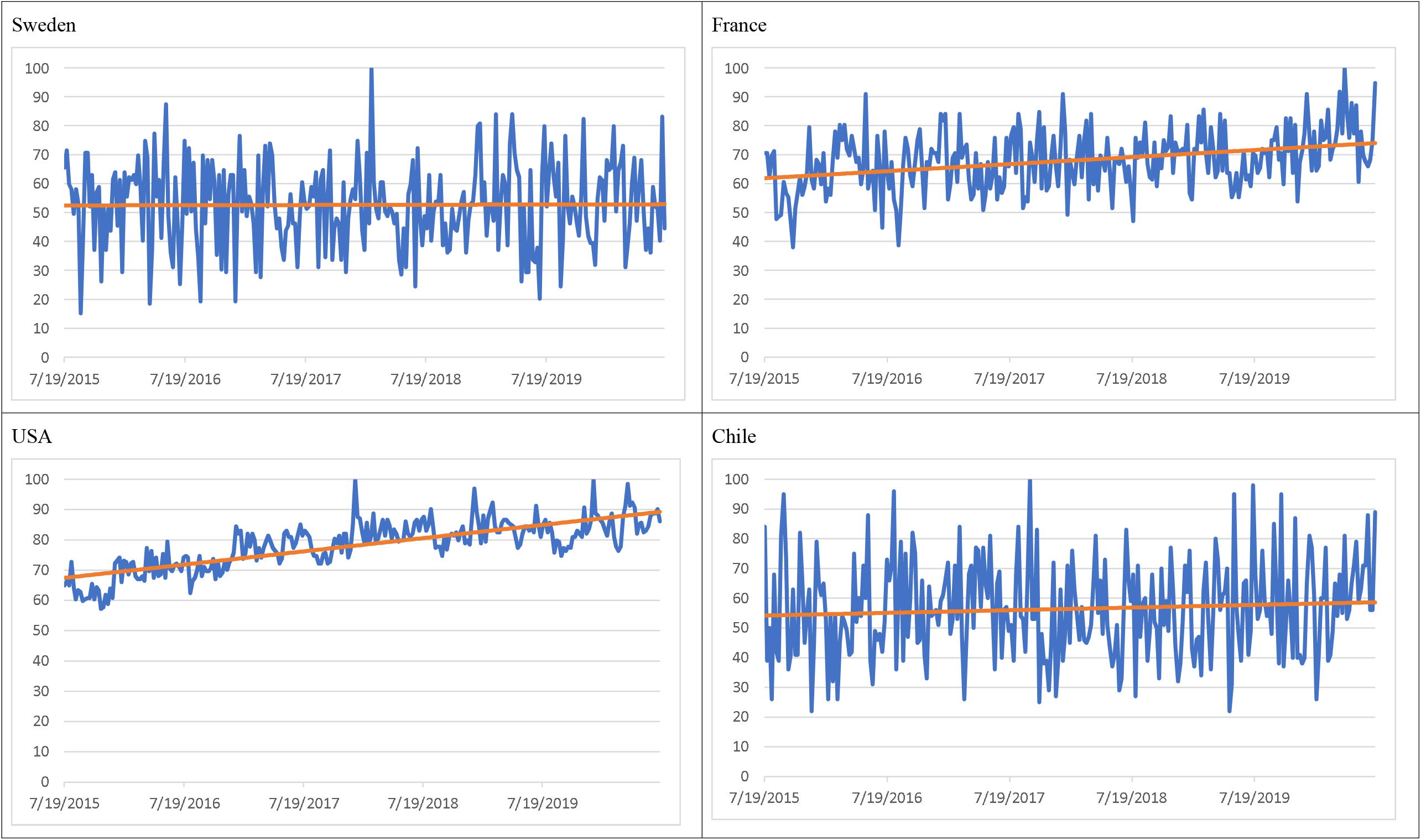
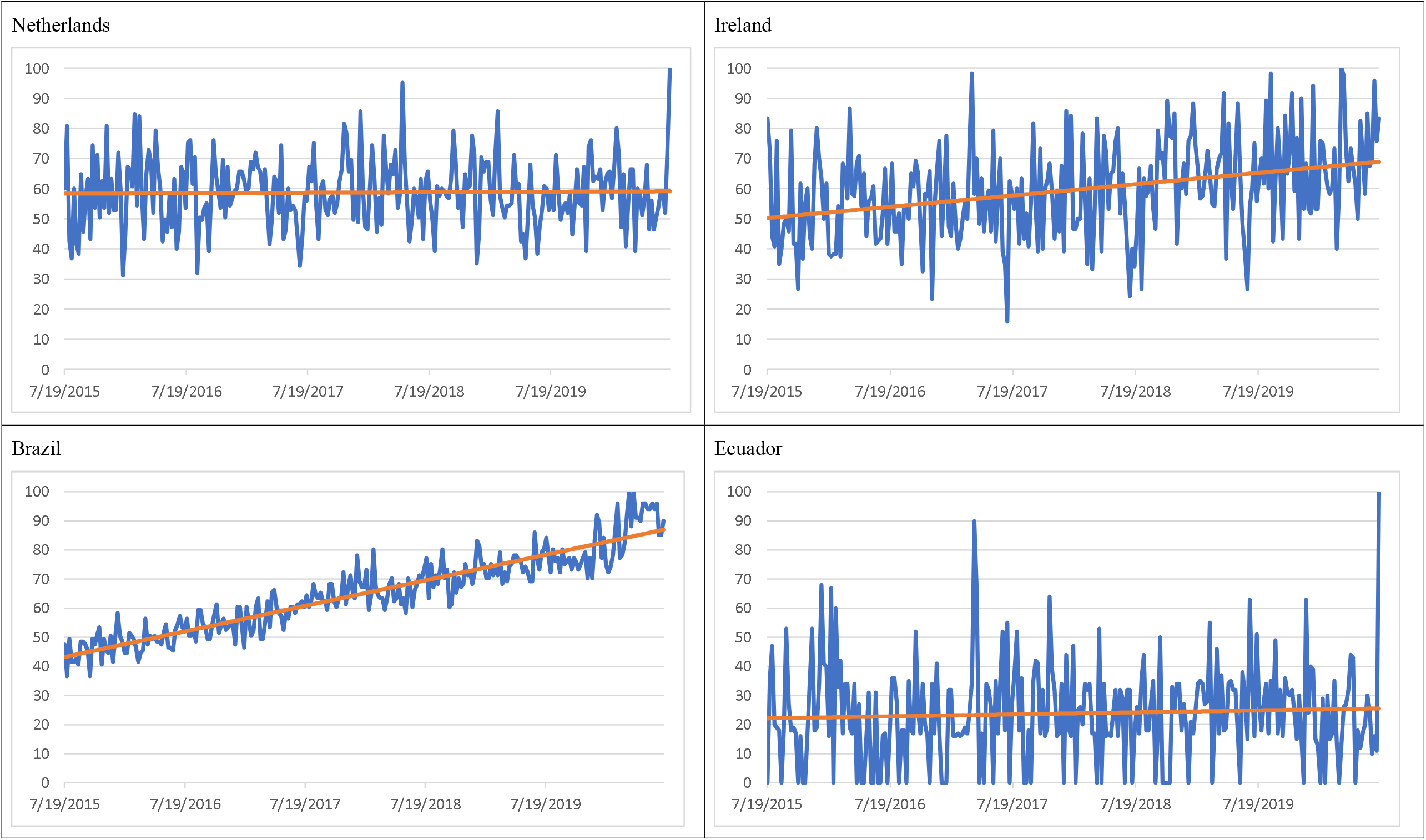
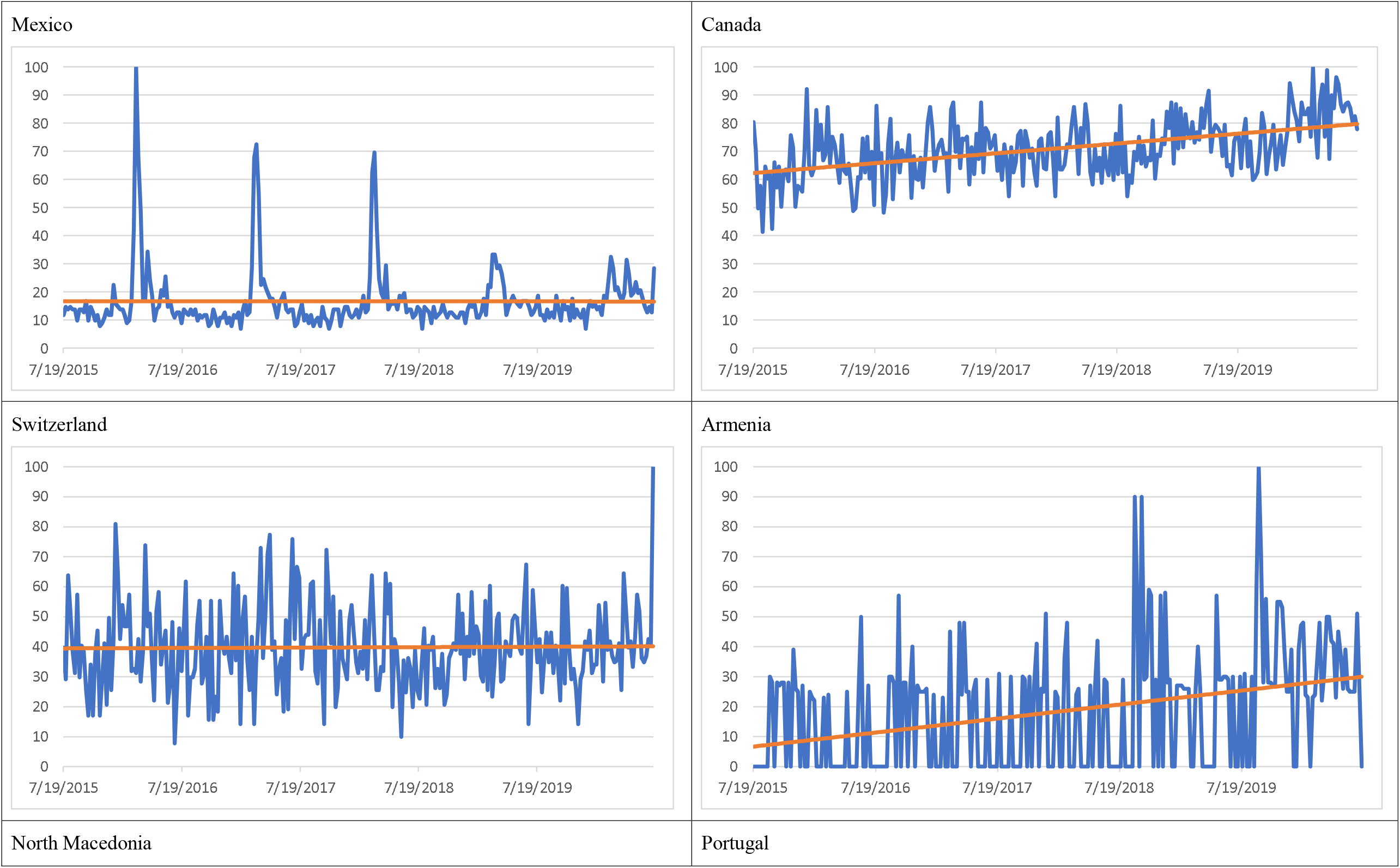
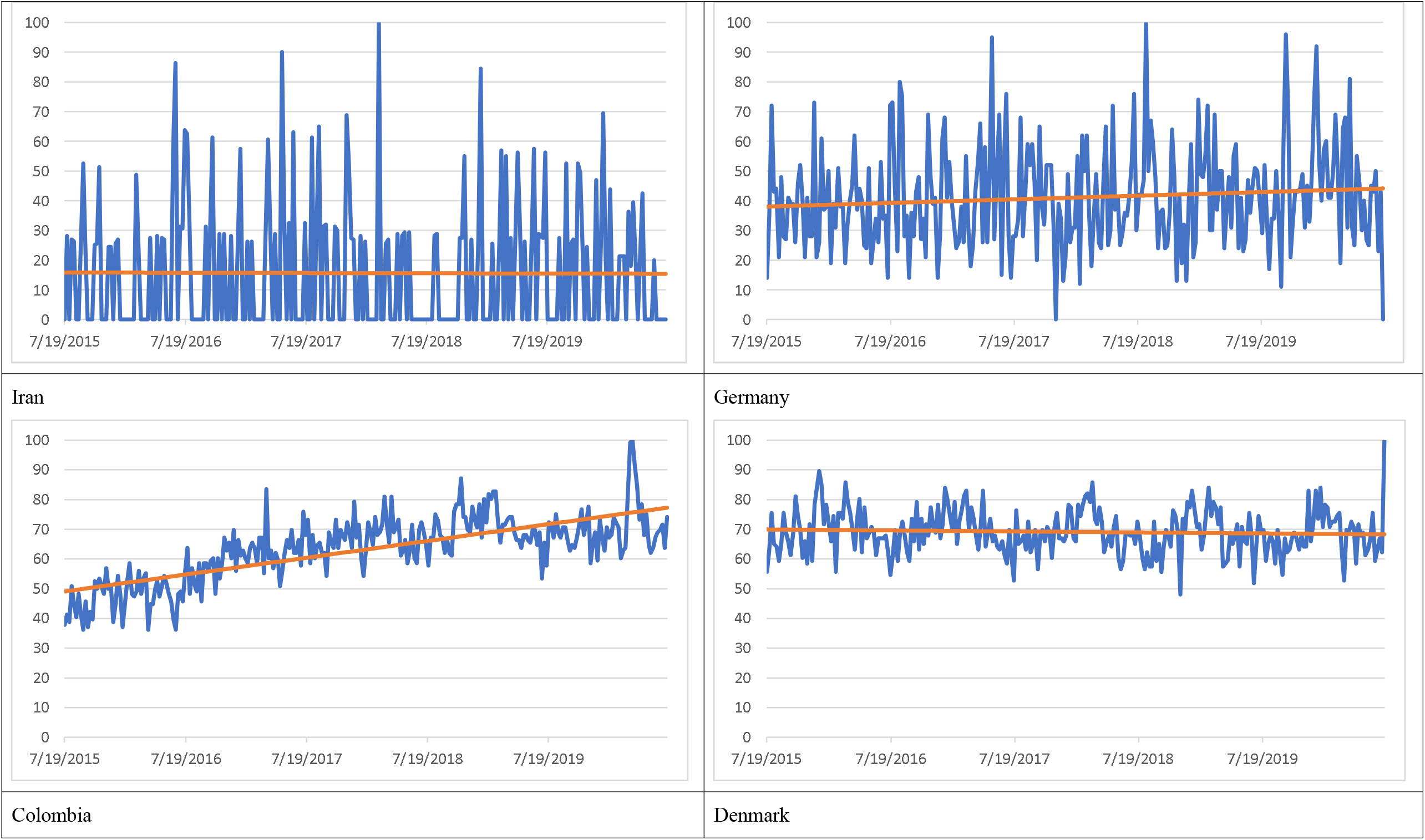
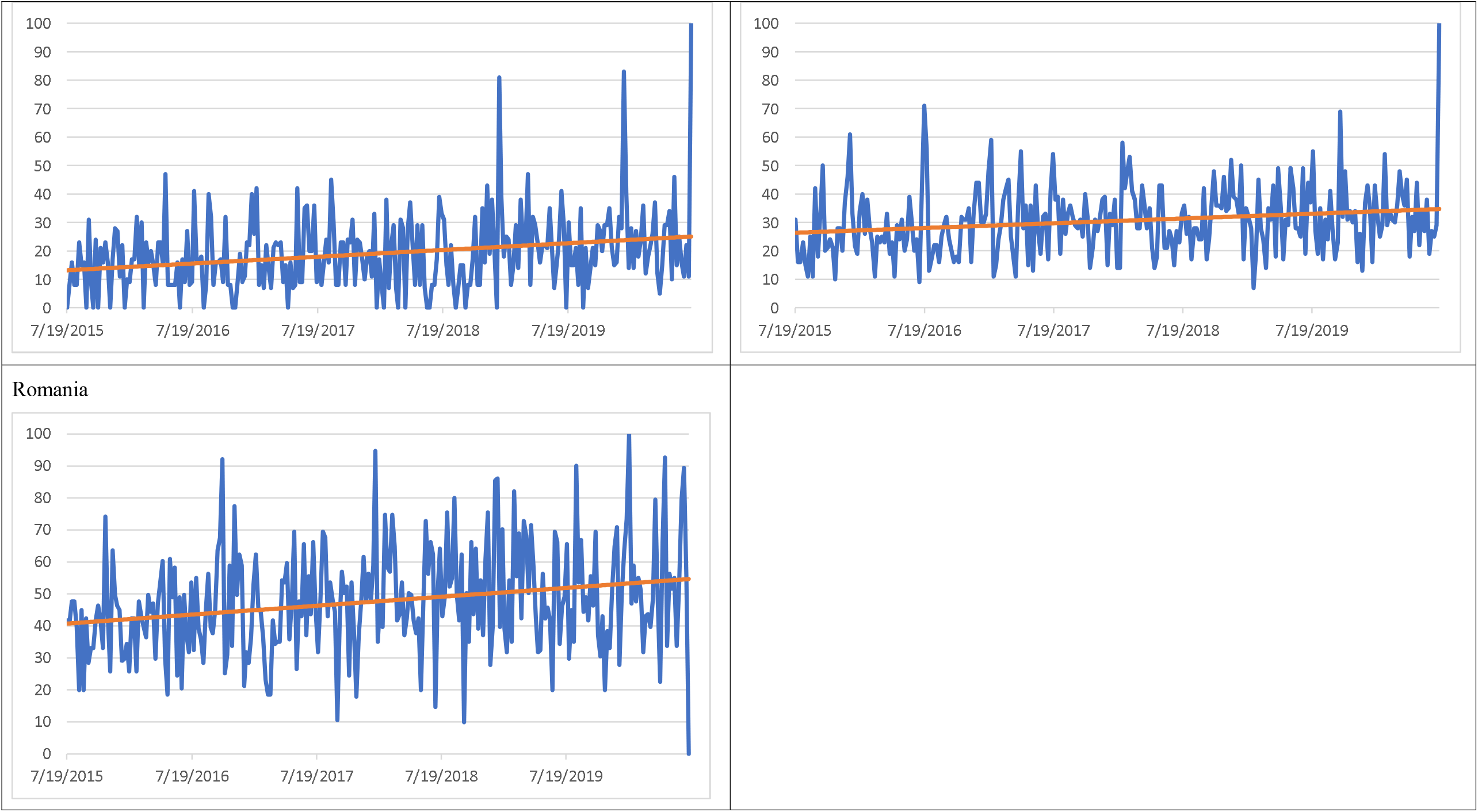

